# Feasibility of SMS-based screening for mood disturbances after minor stroke: strengths and limitations

**DOI:** 10.64898/2026.09.09.26362636

**Authors:** Suhrit Duttagupta, Sylvie Berthoz, Igor Sibon

## Abstract

Stroke frequently causes invisible sequelae such as depression, anxiety, fatigue, and cognitive dysfunction that remain underdiagnosed. Simple and accessible screening tools are needed to support their early detection. This study assessed the feasibility and validity of an SMS-based questionnaire in identifying mood disturbances, fatigue and cognitive complaints before follow-up visits around 3-6 weeks post stroke. 100 minor stroke patients responded to SMS questions on mood, fatigue, and cognition starting from 15 days before their follow-up. Responses were compared with validated scales: Hospital Anxiety and Depression (HAD), Center of Epidemiological Studies-Depression (CES-D), Multidimensional Fatigue Inventory (MFI), and Montreal Cognitive Assessment (MoCA). Associations were examined through group comparisons and correlation analyses. Two-thirds of patients responded to at least one SMS, with lower stroke severity predicting higher adherence. Among responders, 37.8% showed probable depression and 34.07% exhibited probable anxiety based on mood scales, and 29.3% reported depression and 29.4% reported anxiety based on SMS responses. SMS-identified depression and anxiety were strongly associated with higher CES-D and HAD scores (p < 0.001). SMS fatigue intensity correlated moderately with fatigue and mood scores. SMS-identified cognitive complaints were associated with higher MFI Mental fatigue scores (p = 0.008) but not with MoCA scores. SMS-based screening appears feasible and useful for detecting mood disturbances after minor stroke but is less suitable for cognitive screening. Larger, more diverse studies with refined measures are required to validate SMS as a scalable post-stroke follow-up tool.

**Author Summary:** Stroke survivors often experience psychological troubles such as depression, anxiety, and fatigue. While physical impairments may be prioritized by clinicians, these invisible handicaps affect recovery just as much, which is why identifying them quickly and effectively is important. In our study, we selected 100 patients with non-severe strokes who would have a follow-up appointment between 3 and 6 months after stroke. Starting from fifteen days before the follow-up, patients were asked SMS questions if they had specific psychological problems. At the follow-up, patients were given clinical tests to see if their responses matched with the SMS responses they’d given before. This was the case for depression, anxiety, and fatigue, but not so much for cognitive troubles. Overall, our work shows that SMS questionnaires could be used to understand the patient’s mental state after stroke in an easy and accessible way. For patients who find it difficult to come to a hospital for a follow-up, using such surveys could help doctors understand what specific care they need to provide.

## Introduction

Stroke remains a leading cause of disability worldwide and the second-most common cause of death (1). In addition to visible impairments, many survivors experience ‘invisible’ sequelae - depression, anxiety, fatigue, and cognitive dysfunction - that substantially reduce quality of life and hinder recovery (2,3). These conditions are common but frequently underdiagnosed and undertreated (4). Although predictive models based on factors such as age, initial stroke severity, and psychiatric history may identify patients at risk of developing post-stroke neuropsychiatric complications (5), systematic follow-up screening for all patients remains essential. Guidelines recommend conducting multidisciplinary evaluations within 3-6 months (6), but such assessments are resource-intensive and inconsistently implemented due to staffing and time constraints. Digital health tools provide opportunities for remote monitoring (7), but many require smartphone ownership or advanced digital literacy, posing barriers for some stroke survivors. In contrast, short message service (SMS) is widely accessible and user-friendly, making it a promising option for symptom monitoring with minimal technological demands (8).

This pilot study aimed to evaluate whether SMS-based questionnaires can feasibly detect mood disturbances and cognitive complaints in stroke survivors discharged with minor physical disability. We hypothesized that SMS responses would correlate with validated clinical scales and support early identification of at-risk patients.

## Results

100 patients were enrolled with 61 men and a median age of 66.5 (IQR = 20.0). Median NIHSS at admission was 2.5 (IQR 2) and median prestroke and hospital discharge mRS scores were, respectively, 0 (IQR 0) and 1 (IQR 1) suggesting functional independence. Vascular risk factors included hypertension (52.1 %), smoking (41.5 %), dyslipidemia (38.3 %), diabetes (16 %), and obesity (13.8 %).

Ninety-six patients were present at follow-up, although there were missing data for certain evaluations (Table 1). Median NIHSS and mRS scores were 0 (IQR = 0) and 1 (IQR = 1) respectively and 26.9 % of patients had MoCA scores below 24. Prevalence of probable depression was detected in 37.8 % (CES-D) and 18.6 % (HAD-D) while 34.1 % screened positive for anxiety (HAD-A). Fatigue (MFI ≥ 60) was present in 38.2 %, with high prevalence across subdomains: General 55.3 %, Physical 63.2 %, Activity 51.3 %, Mental 46.1 %, and Reduced Motivation 42.1 %.

**Table 1:**
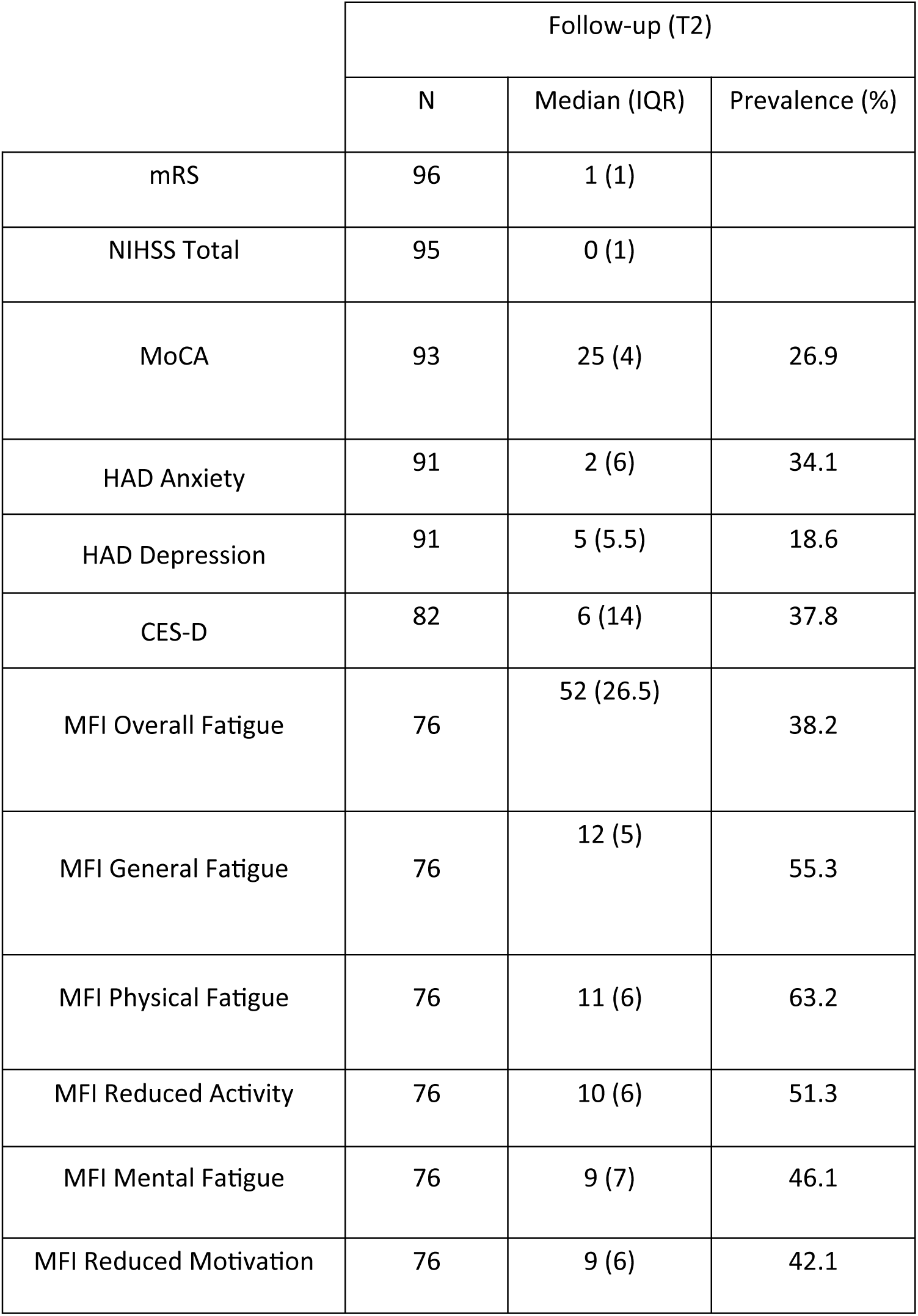
Patient clinical status at follow-up.

Sixty-six patients responded to at least one SMS. Non-responders did not show any significant differences based on stroke risk factors or clinical evaluations with the exception of higher admission NIHSS scores than responders (median: 4 vs 2, p = 0.008).

Table 2 shows the comparison between proportions of groups based on the SMS responses and clinical scale thresholds. For questions on anxiety, fifteen out of fifty-one (29.4 %) responders indicated increased symptoms. The ANX group was significantly more likely to endorse probable anxiety based on the HAD-A (OR: 4.67, p=0.04), probable depression based on the HAD-D (OR: 19.8, p<0.001) as well as the CES-D (OR: 11.3, p<0.001). They were also more likely to show overall fatigue (OR: 5.40, p=0.020) and Reduced Motivation (OR: 5.5, p=0.046). The DEP group consisted of twelve out of forty-one responders (29.3 %) and was significantly more likely to endorse probable depression on the HAD-D (OR: 74.7, p<0.001) and the CES-D (OR: 11.3, p<0.001). The APAT group consisted of six out of thirty-six responders (14 %) who were significantly more likely to show probable anxiety (OR: 43.3, p=0.001) and depression (HAD-D OR: 9.30, p=0.043; CES-D OR: 28.8, p=0.004), Mental fatigue (OR: 18.2, p=0.021) and Reduced Motivation (OR: 14.5, p=0.024). Eleven out of thirty-five responders (24.4 %) indicated cognitive impairment, but there were no significant differences between the COG-IMP and NO-COG-IMP groups based on the clinical scale thresholds.

**Table 2:**
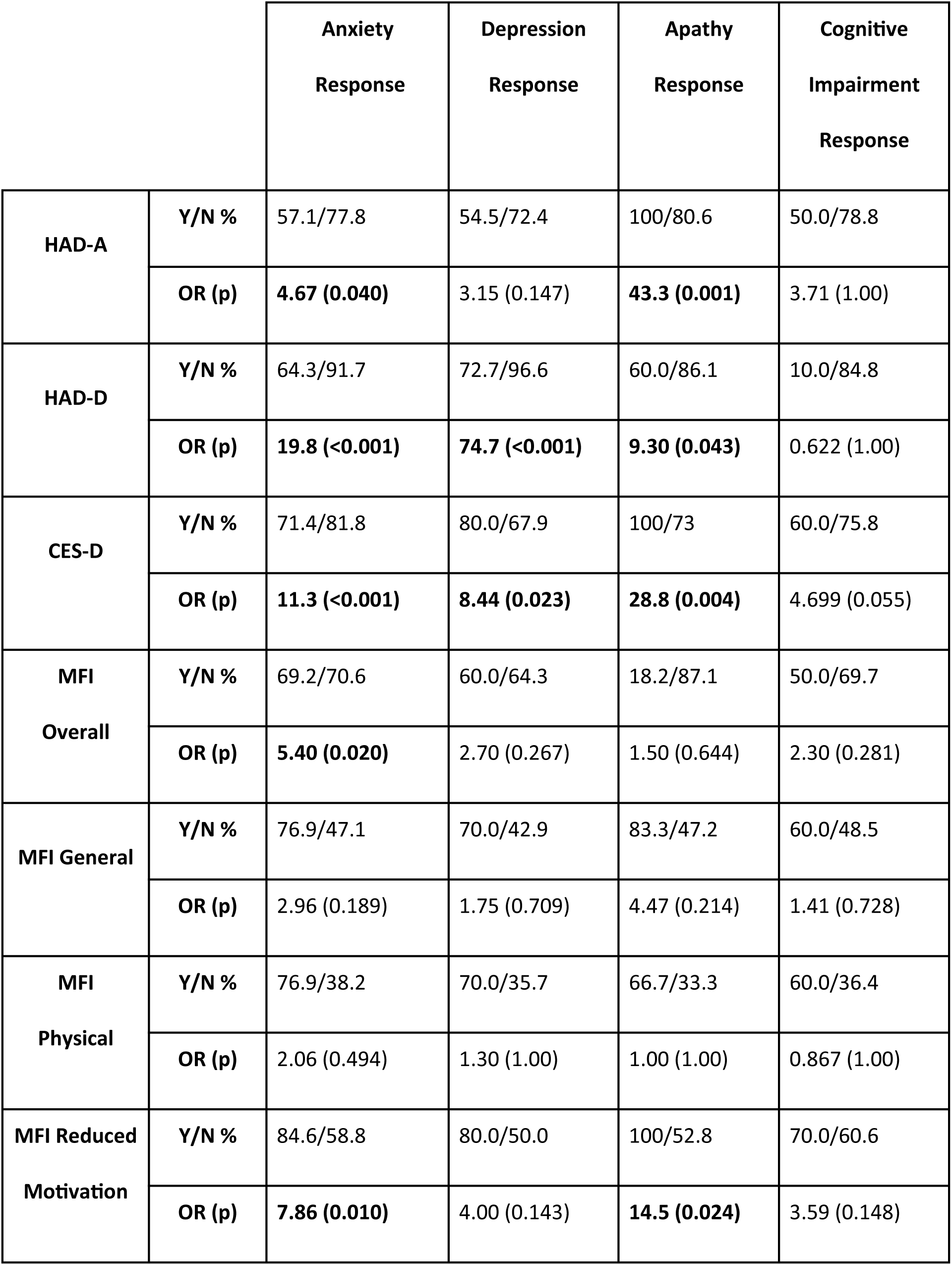
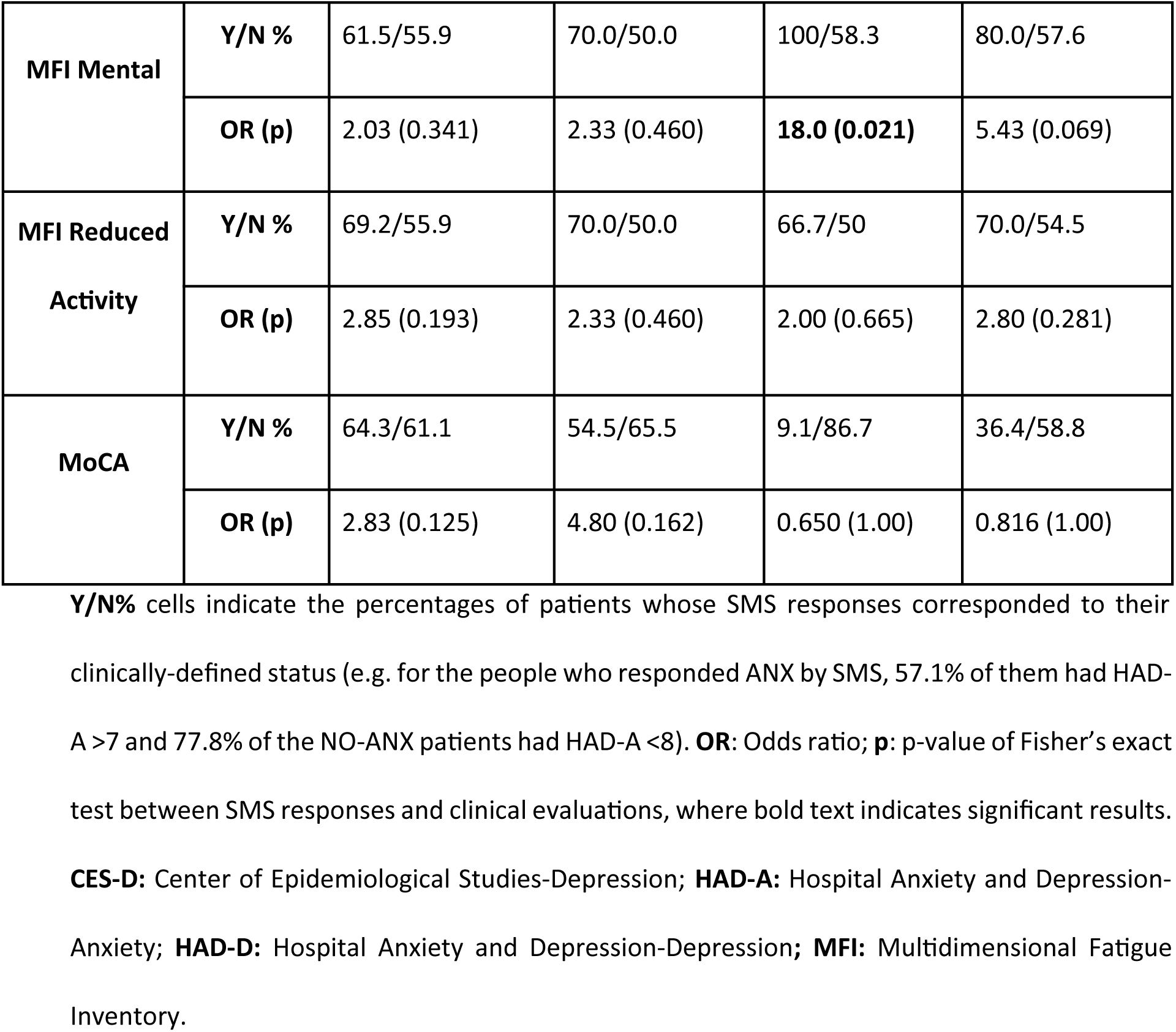
Group comparisons between clinical evaluations and SMS responses.

Table 3 summarizes the dimensional associations between SMS responses and relevant clinical scales’ scores at the follow-up visit. ANX group had significantly higher HAD-A (10.5 vs 5.0, p < 0.001), HAD-D (8.5 vs 3.0, p < 0.001), CES-D (27.5 vs 10.0, p < 0.001), and Overall fatigue scores (p = 0.005). The DEP group showed higher HAD-A (8.0 vs 5.0, p = 0.005), HAD-D (9.0 vs 4.0, p < 0.001), CES-D (24.0 vs 11.5, p < 0.001), and Reduced Motivation scores (12.5 vs 8.5, p = 0.042). The COG-IMP group did not have significantly lower MoCA scores (p = 0.462) but had higher HAD-A (7.0 vs 4.0, p = 0.022) and Mental fatigue (12.5 vs 8.0, p = 0.008) scores.

**Table 3:** Associations between SMS responses and follow-up clinical evaluations.

|  | **ANX** | **NO-ANX** | **DEP** | **NO-DEP** | **APAT** | **NO-APAT** | **COG-IMP** | **NO-COG-IMP** |
| --- | --- | --- | --- | --- | --- | --- | --- | --- |
| **n** | 15 | 36 | 12 | 29 | 6 | 36 | 11 | 34 |
| **HAD-A** | 10.50 | 5.00 | 8 | 5 | 12 | 5 | 7 | 4 |
| **IQR** | (10.8) | (4.00) | (5.50) | (6.00) | (3.00) | (3.00) | (3.75) | (3.00) |
| **Effect (p)** | **0.669 (<0.001)** |  | **0.586 (0.005)** |  | **0.906 (0.001)** |  | **0.482 (0.022)** |  |
| **HAD-D** | 8.50 | 3.00 | 9 | 4 | 8 | 3.5 | 4 | 4 |
| **IQR** | (4.75) | (4.75) | (3.50) | (3.00) | (3.00) | (4.25) | (4.00) | (5.00) |
| **Effect (p)** | **0.704 (<0.001)** |  | **0.806 (<0.001)** |  | **0.756 (0.007)** |  | 0.088 (0.685) |  |
| **CES-D** | 27.5 | 10.0 | 24 | 11.5 | 35 | 11 | 20.5 | 11 |
| **IQR** | (18.5) | (8.00) | (12.8) | (8.25) | (13.0) | (12.0) | (10.0) | (8.00) |
| **Effect (p)** | **0.723 (<0.001)** |  | **0.754 (<0.001)** |  | **0.957 (<0.001)** |  | 0.406 (0.056) |  |
| **MFI Overall** | 65.0 | 47.0 | 62 | 51.5 | 64 | 49 | 56.5 | 47 |
| **IQR** | (17.0) | (29.0) | (32.3) | (24.3) | (7.25) | (28.5) | (15.5) | (30.0) |
| **Effect (p)** | **0.523 (0.006)** |  | 0.371 (0.088) |  | 0.4815 (0.064) |  | 0.333 (0.117) |  |
| **MFI General** | 16 | 11.5 | 15.0 | 12.5 | 15.5 | 11.5 | 12 | 11 |
| **IQR** | (6.00) | (6.25) | (6.50) | (5.25) | (2.50) | (5.25) | (6.25) | (8.00) |
| **Effect (p)** | **0.495 (0.009)** |  | 0.336 (0.121) |  | 0.486 (0.060) |  | 0.112 (0.603) |  |
| **MFI Physical** | 14 | 10.5 | 13.0 | 11.5 | 12 | 12 | 10 | 11 |
| **IQR** | (6.00) | (4.75) | (9.25) | (5.25) | (5.00) | (6.50) | (6.50) | (7.00) |
| **Effect (p)** | **0.396 (0.038)** |  | 0.236 (0.279) |  | 0.023 (0.942) |  | 0.024 (0.920) |  |
| **MFI Reduced Activity** | 13 | 9 | 12.5 | 9.50 | 12 | 9.50 | 11.5 | 9 |
|  | (8.00) | (6.00) | (6.75) | (5.25) | (4.50) | (7.00) | (2.75) | (7.00) |
| **Effect (p)** | **0.410 (0.032)** |  | 0.318 (0.143) |  | 0.148 (0.576) |  | 0.270 (0.204) |  |
| **MFI Mental** | 9.00 | 7.50 | 10.5 | 8.50 | 13 | 7.50 | 12.5 | 8 |
| **IQR** | (6.00) | (8.00) | (5.75) | (6.25) | (4.75) | (6.50) | (4.00) | (5.00) |
| **Effect (p)** | 0.276 (0.147) |  | 0.211 (0.334) |  | **0.620 (0.016)** |  | **0.561 (0.008)** |  |
| **MFI Reduced**<br>**Motivation** | 11 | 8 | 12.5 | 8.50 | 13.5 | 8 | 11 | 8 |
| **IQR** | (5.00) | (5.75) | (4.50) | (5.25) | (3.75) | (5.00) | (5.25) | (5.00) |
| **Effect (p)** | **0.484 (0.011)** |  | **0.439 (0.042)** |  | **0.699 (0.007)** |  | 0.294 (0.165) |  |
| **MoCA** | 25 | 26 | 25 | 26 | 25.5 | 26 | 27 | 26 |
| **IQR** | (3.00) | (4.25) | (3.00) | (5.00) | (2.50) | (4.00) | (3.50) | (3.00) |
| **Effect (p)** | 0.0303 (0.912) |  | 0.191 (0.357) |  | 0.185 (0.478) |  | 0.150 (0.462) |  |

Based on the SMS groups reporting or not reporting difficulties, the median and IQR (parentheses) values of their clinical evaluation scores. The effect size is the rank biserial correlation with p-values in parentheses, bold text indicating significant results. **CES-D**: Center of Epidemiological Studies-Depression; **HAD-A**: Hospital Anxiety and Depression-Anxiety; **HAD-D**: Hospital Anxiety and Depression-Depression; **MFI**: Multidimensional Fatigue Inventory; **MoCA**: Montreal Cognitive Assessment.

Spearman correlations of SMS fatigue intensity with relevant clinical scale scores are presented in Fig 1. Fatigue intensity correlated with Overall fatigue (ρ = 0.417, p = 0.002), HAD-A (ρ = 0.466, p < 0.001), HAD-D (ρ = 0.495, p < 0.001), and CES-D scores (ρ = 0.638, p < 0.001).

**Fig 1.**
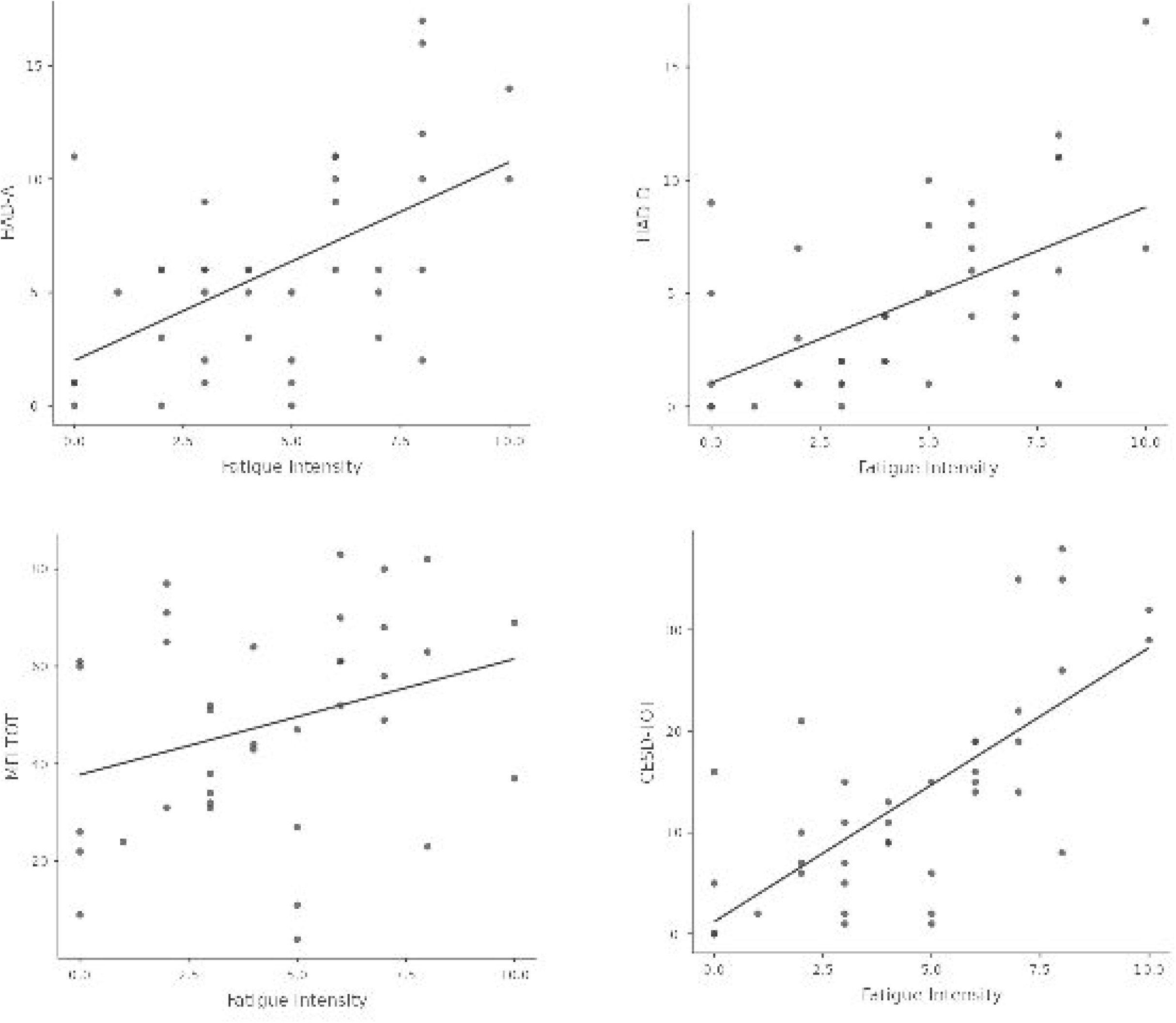
Correlations between SMS fatigue and clinical scales. Scatterplot and linear regression line between the fatigue severity reported by patients through SMS with mood and fatigue evaluations. CES-D: Center of Epidemiological Studies-Depression; HAD-A: Hospital Anxiety and Depression-Anxiety; HAD-D: Hospital Anxiety and Depression-Depression; MFI-TOT: Multidimensional Fatigue Inventory Overall fatigue.

**Fig 2.**
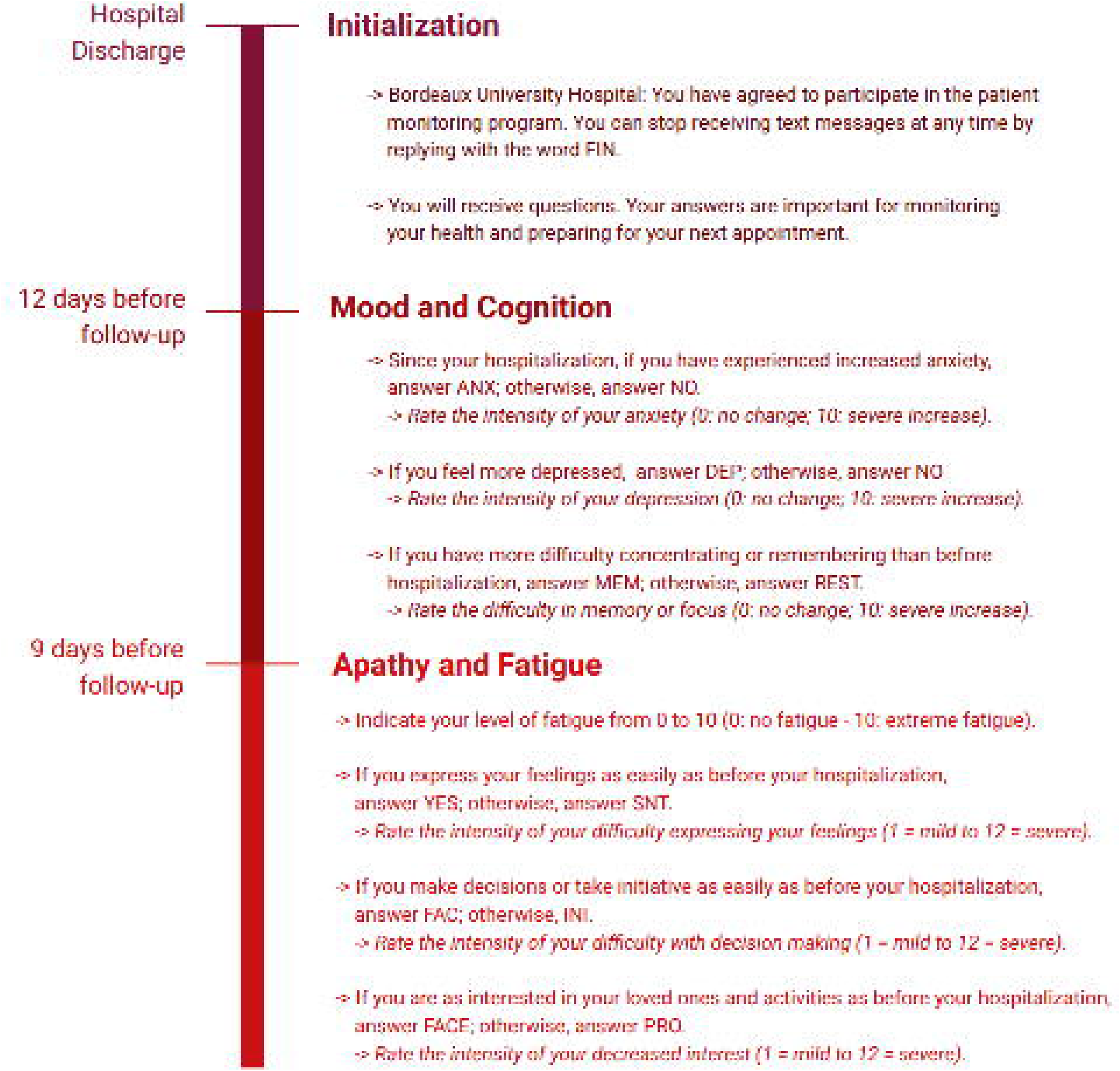
SMS follow-up survey. Timeline of questions sent to patients via SMS. Questions in italics were only asked if the prior response indicated the presence of difficulty.

## Discussion

This study demonstrates the feasibility of SMS-based screening for mood disturbances after minor stroke. Two-thirds of patients responded, where (i) SMS-identified anxiety and depression correlated strongly with validated scales, (ii) responses for fatigue were associated with the corresponding clinical evaluation as well as emotional impairments, and (iii) SMS-reported cognitive complaints were not associated with clinical MoCA scores. Our results illustrate how simple, low-barrier digital tools can detect at-risk individuals before follow-up visits.

These results are in line with a recent review from Shalaby et al. demonstrating the effectiveness and patient satisfaction of text message evaluations in the field of mental health (15). They confirm the utility of mobile technology in the early identification of post-stroke patients (16,17) who may benefit from a comprehensive psychiatric evaluation and corresponding care pathway. Interestingly, we observed a discrepancy between the depression measures: while both HAD-D and CES-D identified depressive symptoms, CES-D showed higher sensitivity and stronger associations with SMS reports. This confirms the importance of using CES-D in post-stroke populations compared to shorter instruments such as the HAD-D due to its multidimensionality (18). Regarding cognition, the absence of an association between SMS-reported complaints and MoCA scores deserves attention. It must be noted that out of the eleven patients reporting cognitive difficulties by SMS, only five had a recorded MoCA score. Nevertheless, several mechanisms may explain this discrepancy. First, the restricted variability of MoCA in a relatively mildly impaired population likely reduced statistical sensitivity. Second, subjective cognitive complaints are often influenced by mood disorders, with anxiety and depression heightening perceived cognitive dysfunction even when objective tests indicate otherwise (19). Thus, even if SMS cognitive items may reflect subjective distress rather than neurocognitive impairment, they may signal psychological burden requiring targeted intervention and therefore remain clinically relevant. Moreover, these discrepancies suggest that subtle cognitive impairments observed in minor stroke patients may require more comprehensive neuropsychological testing (20).

Beyond feasibility, these findings provide insight into the interplay between mood, cognition, and fatigue in stroke survivors. Anxiety and depression were not only prevalent but also closely associated with fatigue scores, supporting prior evidence that mood disturbances amplify the subjective burden of fatigue (21). The moderate correlations between SMS fatigue ratings and CES-D/HAD-A further suggest that fatigue reports captured by SMS may partly reflect underlying mood symptoms rather than purely physical exhaustion. These results also reinforce the hypothesis that post-stroke fatigue is complex and multifaceted, likely exacerbated by mood problems such as high depressivity and anxiety (22). Similarly, cognitive impairments can heighten the perception of fatigue, creating a feedback loop that amplifies both fatigue and mood-related symptoms. Understanding these links can aid clinicians in tailoring interventions to target specific dimensions of fatigue, ultimately improving both the psychological and cognitive recovery of stroke patients. Overall, the use of multidimensional fatigue assessments is warranted since unidimensional scales or single-item ratings may underestimate the complexity of post-stroke fatigue.

Several limitations constrain interpretation: (i) the study cohort largely consisted of patients with mild strokes, restricting generalizability to more severe stroke populations, (ii) a high non-response rate (34 %) introduces potential selection bias because patients with greater baseline disability were less likely to engage, (iii) relying solely on the MoCA may have underestimated subtle cognitive deficits, (iv) missing data and absence of adjustments for multiple comparisons raise the risk of false-positive associations, (v) relevant comorbidities were not systematically assessed, and neither patients’ nor providers’ perceptions of SMS usability were captured. Finally, (vi) conceptual overlap between SMS questions and clinical scales may have inflated correlations.

In conclusion, our study highlights the use of SMS as a pragmatic, scalable screening tool for mood disturbances in stroke survivors, particularly in older adults who may struggle with more complex digital platforms. By identifying patients at risk before follow-up visits, SMS-based tools could enhance triage, support early intervention, and reduce underdiagnosis of post-stroke depression and anxiety. To realize this potential, future studies must recruit more heterogeneous cohorts, conduct refined neurocognitive and psychiatric assessments, evaluate acceptability from user perspectives, and implement strategies to improve adherence.

## Methods

### Study Design and Participants

Patients admitted to the Bordeaux University Hospital stroke unit between 01/12/2022 and 31/08/2024 were screened for the study and data were collected from the institution’s electronic database. Oral consent was obtained, as documented by the attending nurse and witnessed by the neurologist, to receive SMS messages starting from 15 days before follow-up visits; written consent was not required since all investigations were carried out as part of standard clinical practice. Sending text messages to collect information prior to the follow-up visit was already in use at our institution as a routine procedure to monitor patients following orthopaedic surgery. The full protocol was approved by our institutional ethics committee (rCER-BDX 2024-75)

The main inclusion criteria were: (i) stroke confirmed on brain MRI, (ii) discharged home after admission to the stroke unit, (iii) scheduled for follow-up 3 to 6 months post-stroke, and (iv) ability to use a mobile phone for SMS. The main exclusion criteria were motor, speech, or cognitive disability judged too severe for SMS communication.

### Data Collection

Data were accessed for research purposes on 05/11/2024. Baseline demographics, vascular risk factors, and stroke characteristics were extracted from the electronic medical record completed at baseline and post-stroke visits. Stroke severity was measured with the National Institute of Health Stroke Scale (NIHSS) (9) and functional outcome with the modified Rankin Scale (mRS) (10).

Cognitive and emotional impairments were assessed by the Montreal Cognitive Assessment (MoCA) (11), Center of Epidemiologic Studies-Depression (CES-D) (12), and Hospital Anxiety and Depression (HAD) scales (12). A MoCA score < 24 was considered indicative of cognitive impairment. A CESD-D score ≥ 16 or an HAD-D score ≥ 8 were considered indicative of probable depression while a HAD-A score ≥ 8 was considered suggestive of anxiety. Fatigue was assessed using the Multidimensional Fatigue Inventory (MFI), which covers five dimensions of fatigue: General fatigue, Physical fatigue, Reduced Motivation, Reduced Activity, and Mental fatigue (13), with a total score ≥ 60 indicating Overall fatigue. The following cut-offs have been recommended for the subscores: 11 for General fatigue, 10 for Reduced Activity and 9 for Physical fatigue, Reduced Motivation and Mental fatigue each (14).

### SMS Questionnaire

The SMS follow-up protocol used three series of questions to interview patients regarding their experiences of anxiety, depression, attention and memory difficulties, fatigue, and apathy (identified as problems with emotional self-expression, decision-making difficulties and reduced activity following stroke) and spasticity. To improve adherence to the protocol, the three sets of questions were sent on separate days before the follow-up visit: spasticity and pain 15 days prior; mood and cognition 12 days prior; fatigue and apathy 9 days prior (Figure 1). For the present study, we did not use the responses for spasticity and pain because patients had low physical disability at hospital discharge.

## Statistical Analysis

Descriptive analyses were performed with numeric variables expressed as median (IQR) and discrete variables as absolute and relative frequencies (%). Four groups were created according to emotional and cognitive SMS responses (Yes/No): Anxiety (ANX vs NO-ANX), Depression (DEP vs NO-DEP), Apathy (APAT vs NO-APAT), and Cognitive Impairment (COG-IMP vs NO-COG-IMP). Group comparability was tested for age and sex.

All analyses were performed with respect to group comparability. Continuous values were compared using the Mann-Whitney U-test and discrete results were compared using chi-square or Fisher’s exact test. The alpha risk was set at 5 % and two-tailed tests were used.

Correlations between SMS fatigue intensity evaluation and MFI, HAD-A, HAD-D, and CES-D scores were computed using Spearman’s coefficients (ρ).

Since the analyses were exploratory, no adjustment for multiple comparisons was applied. No missing data was imputed. Statistical analysis was performed with Jamovi (version 2.6, www.jamovi.org).

## Data Availability

All data produced in the present study are available upon reasonable request to the authors.

## Acknowledgements

The authors would like to thank the French company Calmedica for the use of their tool, memoQuest, in the study.

